# Altered association of plasmatic oxytocin with affective stress response in alcohol use disorder

**DOI:** 10.1101/2024.09.30.24314609

**Authors:** Annalina V. Mayer, Yana Schwarze, Janine Stierand, Johanna Voges, Alexander Schröder, Janina von der Gablentz, Klaus Junghanns, Oliver Voß, Sören Krach, Frieder M. Paulus, Lena Rademacher

## Abstract

Oxytocin has been proposed to play a role in the development and maintenance of alcohol use disorder (AUD) through its interactions with stress pathways. Empirical evidence that indicates altered associations between endogenous oxytocin and stress reactivity in AUD is currently lacking. In this study, we investigated baseline plasmatic oxytocin concentrations of early-abstinent patients with AUD (*N* = 40) and matched healthy control participants (*N* = 37), who completed the Trier Social Stress Test (TSST) as well as a control task on two separate visits. We measured salivary cortisol and pulse rate as indicators of a physiological stress response, and anxiety ratings as an indicator of an affective stress response at multiple time points. Baseline oxytocin levels did not significantly differ between the groups. However, our results suggest an altered association of oxytocin and affective stress responses in participants with AUD: while participants with AUD showed a positive relationship between plasmatic oxytocin levels and stress-induced anxiety increase, the opposite relationship was found in control participants. We did not find evidence for an association of oxytocin with physiological stress responses. Assuming that affective stress responses mediate addiction-related behaviors like craving and relapse, our findings suggest that administering exogenous oxytocin might not be beneficial in treating AUD, at least during the early stages of abstinence.

## Introduction

Various studies have highlighted the importance of stress in the development and maintenance of alcohol use disorder (AUD) (Anthenelli, 2012). The relationship between heavy alcohol consumption and stress seems to be reciprocal: while repeated exposure to stressful situations is an important risk factor for AUD and increases the risk of relapse (Keyes et al., 2011; Milivojevic & Sinha, 2018), chronic alcohol misuse itself stimulates the release of major stress hormones, including corticotropin-releasing factor (CRF), adrenocorticotropin (ATCH) and cortisol (Rivier & Lee, 1996). In the long run, the body’s chronic efforts to adapt to repeated or prolonged stress lead to a cumulative physiological toll on the individual, which has been referred to as allostatic load (McEwen & Stellar, 1993). In line with this, attenuated physiological stress responses have been observed in individuals with AUD, suggesting a dysregulation of the hypothalamic–pituitary–adrenal (HPA)-axis and the autonomic nervous system after prolonged periods of heavy alcohol use (Boschloo et al., 2011; Chen et al., 2020; Dunne & Ivers, 2023; Junghanns et al., 2003, 2005; Lovallo et al., 2000; Sinha et al., 2009). Notably, while physiological stress responses are blunted, affective stress responses such as feelings of anxiety, worry and unease seem to be comparable or even more pronounced in individuals with AUD relative to healthy individuals, even in long-term abstinence (Romero-Martínez et al., 2019; Sinha et al., 2009; Starcke et al., 2013).

Recently, the hormone and neuropeptide oxytocin has become a focus of research on the link between stress and addiction. A large body of research has discussed the role of oxytocin in modulating stress responses (Love, 2018; Matsushita et al., 2019). Newer frameworks characterize oxytocin as an allostatic hormone, implying that it facilitates adaptation to anticipated changes in the environment and that alterations in the oxytocin system might be associated with dysregulated stress responses (Quintana & Guastella, 2020; Takayanagi & Onaka, 2022). For example, results from rodent studies show that inhibiting the action of endogenous oxytocin in the brain through administration of an oxytocin receptor antagonist results in increased basal and stress-induced activity of the HPA-axis (Hodges et al., 2019; Neumann et al., 2000). In humans, converging evidence suggests that plasmatic and salivary oxytocin levels rapidly increase during and after stressful conditions (Bernhard et al., 2018; Jong et al., 2015; Pierrehumbert et al., 2010; Sanders et al., 1990). This short-term increase is thought to dampen physiological stress responses by modulating HPA-axis activity (Winter & Jurek, 2019). In line with these findings, higher overall plasma oxytocin levels were associated with dampened physiological responses to different stressors in postpartum mothers (Grewen & Light, 2011). Findings on the relationship of oxytocin and affective stress responses, however, have been mixed (Takayanagi & Onaka, 2022). Although some studies suggest that oxytocin may facilitate acute fear responses, most studies report an anxiolytic effect of oxytocin in healthy individuals (Janeček & Dabrowska, 2019), suggesting lower levels of anxiety in those with higher endogenous oxytocin levels (Carson et al., 2015; Heinrichs et al., 2003; Weisman et al., 2013) and after intranasal oxytocin administration (de Oliveira, Chagas, et al., 2012; de Oliveira, Zuardi, et al., 2012).

Aside from its involvement in stress regulation, oxytocin has also been suggested to modulate alcohol-related responses, such as craving (Hansson et al., 2018; McGregor & Bowen, 2012), tolerance (Szabó et al., 1985), and withdrawal symptoms (Szabó et al., 1987). Consequently, it has been proposed that alcohol and other substance use disorders are characterized by alterations in the oxytocin system (King et al., 2020; Vaht et al., 2016). This hypothesis is supported by evidence from animal models of AUD (e.g., Sarnyai & Kovács, 1994; Stevenson et al., 2017), human studies linking alcohol consumption to plasma oxytocin levels and DNA methylation in the oxytocin receptor gene (Rung et al., 2022), as well as clinical studies showing elevated plasma oxytocin levels in patients with AUD during the first days of abstinence compared to healthy individuals (Lenz et al., 2021; Marchesi et al., 1997).

It has been proposed that oxytocin may exert its effects on addiction by interacting with stress pathways (Bowen et al., 2016). For example, central administration of oxytocin or the oxytocin analog carbetocin was found to inhibit drug-seeking behavior after stress in rodents (Qi et al., 2009; Zanos et al., 2014). In humans, intranasal oxytocin was similarly found to decrease stress-induced anxiety and craving of cannabis in a randomized controlled trial (McRae-Clark et al., 2013). These findings suggest that by dampening stress responses, both affective and physiological, oxytocin may facilitate abstinence (Bowen et al., 2016). It has been argued that this makes the oxytocin system a potential pharmacological target for treatments of substance use disorders (Lee & Weerts, 2016). However, although preclinical studies on the effects of exogenous oxytocin appear promising, no research has demonstrated a direct connection between altered endogenous oxytocin levels and stress reactivity in individuals with substance use disorders, including AUD. In AUD specifically, it seems to be relevant to distinguish between affective and physiological stress responses, since these are differently affected by prolonged periods of heavy alcohol consumption (Sinha et al., 2009).

The aim of the present study was thus to examine associations between plasmatic oxytocin levels with affective and physiological responses to psychosocial stress. To test this, participants with and without AUD underwent the Trier Social Stress Test (TSST), a validated task that reliably induces stress, as well as a control task lacking the elements of stressful social evaluation. Salivary cortisol samples, pulse rate, and affect ratings were collected before, during and after the tasks and their association with baseline plasmatic oxytocin levels was examined. For healthy control participants, based on research showing anxiolytic effects of oxytocin, we expected lower stress responses in individuals with higher baseline oxytocin levels. Since participants with AUD might show a different pattern of oxytocin-related stress responses compared to controls, we examined interaction effects of group, task and oxytocin on physiological and affective stress measures.

## Methods

### Participants

Forty-two participants with a DSM-5 diagnosis of moderate or severe alcohol use disorder (fulfilling 5 to 11 criteria) and 38 healthy control participants between the ages of 24 and 59 years were recruited. Two participants with AUD and one control participant discontinued the study, leaving data from 77 participants to be analyzed (see Table 1 for sample characteristics). Participants were enrolled at the Center of Brain, Behavior and Metabolism in Lübeck, Germany between January 2019 and October 2022, with a break due to restrictions during the SARS-CoV-2 pandemic between March 2020 and October 2021.

**Table 1:** Sample characteristics.

|  | AUD (N=40) | Control group (N=37) | Total (N=77) | group comparison<br>p-value |
| --- | --- | --- | --- | --- |
| **Gender** |  |  |  |  |
| female | 2 (5.0%) | 2 (5.4%) | 4 (5.2%) |  |
| male | 38 (95.0%) | 35 (94.6%) | 73 (94.8%) |  |
| **Age [years]** |  |  |  |  |
| Mean (SD) | 42.23 (9.15) | 43.38 (9.68) | 42.78 (9.36) |  |
| Range | 24 - 59 | 25 - 59 | 24 - 59 |  |
| **Body mass index [kg/m<sup>2</sup>]** |  |  |  | 0.042 |
| Mean (SD) | 24.84 (3.56) | 26.86 (4.95) | 25.81 (4.38) |  |
| Range | 16.90 - 30.90 | 20.28 - 44.30 | 16.90 - 44.30 |  |
| **Depressive symptoms [BDI-FS]** |  |  |  | < 0.001 |
| N-Miss | 0 | 1 | 1 |  |
| Mean (SD) | 3.79 (2.97) | 1.05 (1.33) | 2.49 (2.71) |  |
| Range | 0 - 12 | 0 - 5 | 0 - 12 |  |
| **Trait stress reactivity [SRS]** |  |  |  | < 0.001 |
| Mean (SD) | 56.32 (10.62) | 47.22 (7.80) | 51.95 (10.37) |  |
| Range | 36 - 81 | 36 - 67 | 36 - 81 |  |
| **Chronic stress [TICS]** |  |  |  | < 0.001 |
| Mean (SD) | 21.17 (8.00) | 10.68 (5.12) | 16.13 (8.55) |  |
| Range | 2 - 36 | 3 - 24 | 2 - 36 |  |

Note. BDI-FS: Beck Depression Inventory Fast Screen (Beck et al., 2000), SRS: Stress-Reactivity-Scale (Schulz et al., 2005), TICS: screening scale of the Trier Inventory for the Assessment of Chronic Stress (Schulz & Schlotz, 1999). *p*-values derive from group comparisons using two-sample *t*-tests.

Participants with AUD were recruited from addiction wards of two local inpatient psychiatric clinics. All had successfully completed physical withdrawal and had been abstinent for 9 to 39 days at the time of study participation (*M* = 14.3, median = 12, *SD* = 6.3 days). Control participants were recruited by public notices, online advertisements and by letter via the residents’ registration office. The control group was matched to the AUD group in terms of age and gender. All inclusion and exclusion criteria as well as changes to the study protocol due to the SARS-CoV-2 pandemic are listed in the supplementary materials. This study was approved by the Ethics Committee of the University of Lübeck, Germany (AZ 17-077). Full informed written consent was obtained from all participants.

### Study design and procedure

The present analysis was part of a larger functional magnetic resonance imaging (fMRI) study examining acute behavioral, physiological and neural responses to social stress in AUD. Findings on differential stress responses and associated functional connectivity from this sample have been published elsewhere (Schwarze et al., 2024). After an initial screening process ensuring that all inclusion and no exclusion criteria were met, eligible participants were invited to two study visits. For all except for one control participant, these two visits were scheduled between 1 and 9 days apart (*M* = 2.59, *SD* = 2.01). For one control participant, the second visit had to be postponed due to indisposition, which resulted in it not taking place until 48 days after the first visit. Excluding this participant from the analyses did not affect the pattern of results.

Except for an experimental manipulation of social stress, the two visits were identical in procedure (**Figure 1A**). The order of the stress and control day was counterbalanced across participants. At the beginning of each visit, it was ensured that all inclusion criteria were still met, which was then followed by the collection of a first saliva and blood sample.

**Figure 1:**
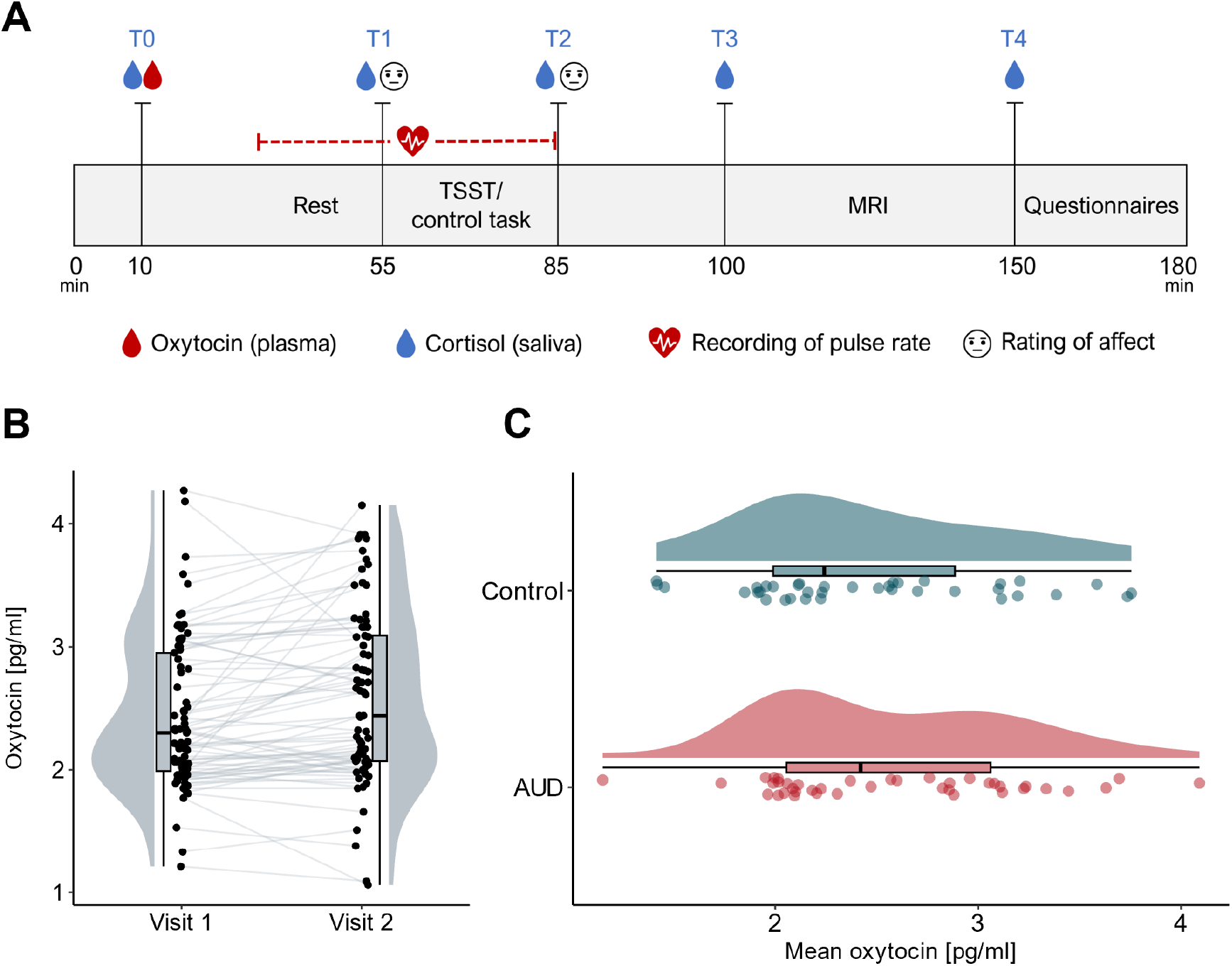
Overview of study design and oxytocin levels. **A:** Course of the two study visits. The visits were identical in procedure except for an experimental manipulation of stress: participants either completed the Trier Social Stress Test (TSST) or a control task. **B:** Within-subject variation of plasmatic oxytocin levels across two visits (jittered raw data from all participants, boxplots and probability distributions). **C:** Averaged plasmatic oxytocin levels in participants with AUD and control participants (from top to bottom: probability distribution, boxplot, and jittered raw data for participants of each group).

Subsequently, participants were introduced to the tasks they were to perform in the scanner and screened for recent MRI contraindications. After this, participants were told to rest while they were shown a video of landscape scenes for about 20 to 30 minutes. During this period, a pulse oximeter (PULOX PO-300, Novidion GmbH) was attached to the participant’s index finger of the non-dominant hand and pulse rate recordings were started. After the resting period, participants were asked to give a second saliva sample and rate their current level of anxiety using the state scale of the State-Trait Anxiety Inventory (STAI-s) (Grimm, 2009).

They were then introduced to the stress or control task, which took 20 minutes to complete (for details, see below). After the task, pulse rate recordings were stopped and participants were again asked to provide a saliva sample and rate their level of anxiety. Another saliva sample was retrieved about 20 minutes later, which was immediately before participants entered the MRI (for details, see supplementary materials). Afterwards, another saliva sample was collected and participants filled out questionnaires assessing participant health and experienced chronic stress (for details, see below). Both visits lasted around 3 hours in total and were conducted at the same time of day (8:30-11:30 a.m. ± 30 min.) to prevent day time fluctuations in the endocrinological parameters. Participants were advised to abstain from caffeine and exercise in the morning before the visits. Smoking was allowed up until 30 minutes before study participation.

### Stress and control task

The Trier Social Stress Test (Kirschbaum et al., 1993) was used to induce psychosocial stress. The TSST is a validated and widely used paradigm to assess the impact of stress under experimental conditions (Narvaez Linares et al., 2020) and follows a standardized protocol, which consists of a ten-minute anticipation phase and a ten-minute test phase. By placing participants in a situation of social-evaluative threat, the TSST has been shown to produce reliable physiological stress responses such as increased heart rate and cortisol release (Dickerson & Kemeny, 2004). Before the task started, participants were informed that their verbal and mathematical skills would be evaluated and that they would have to perform a short speech in front of a jury. In the anticipation phase, they had 10 minutes to prepare this speech, which they delivered in front of the jury in the subsequent test phase.

Additionally, they were asked to perform a mental arithmetic task.

As a control condition, participants were asked to read a text aloud and solve easy arithmetic tasks in the absence of a jury, ostensibly to measure the physiological correlates of mental effort. During the anticipation phase of the control task, participants were instructed to read through the text. The TSST and the control task took place in the same rooms and participants were asked to stand during the test phase in both conditions. Thus, the control task was comparable to the TSST in terms of physical demands, but lacked the element of social evaluation.

### Cortisol and oxytocin samples

Salivary cortisol was measured from samples collected at five timepoints during the two visits (**Figure 1A**). Saliva was collected using a cotton roll (Neutral Salivettes®, SARSTEDT, Nümbrecht, Germany) and stored in a freezer at −20°C until it was sent to an external laboratory for analysis (Dresden LabService GmbH, Dresden, Germany). The samples were thawed and centrifuged at 3000 rpm for five minutes. Salivary cortisol levels were then determined using a chemiluminescence immunoassay procedure (IBL International GmbH, Hamburg, Germany). The intra- and inter-assay coeficients of variance were below 9%.

Baseline oxytocin levels were determined from blood samples taken at the beginning of each visit. The blood was collected into precooled 2.6 ml S-Monovettes® (SARSTEDT, Nümbrecht, Germany) and then centrifuged for 15 minutes at 4°C and 1000 rpm. The obtained blood plasma was subsequently stored at -80°C until the samples were analyzed by a third party (RIAgnosis, Sinzing, Germany) using a combination of selective extraction and extremely sensitive and specific radioimmunoassay (RIA). This standardized and validated procedure has frequently been used to quantify oxytocin in biological fluids including plasma, saliva, and cerebrospinal fluid (e.g., Jong et al., 2015; Martin et al., 2018; Martins et al., 2020). In brief, plasma samples (0.5ml) were kept at −20°C until extraction using LiChroprep® Si60 (Merck) heat-activated at 690°C for 3h. 20mg of LiChroprep® Si60 in 1ml distilled water were added to the sample, mixed for 30min, washed twice with distilled water and 0.01mol/l HCl and eluded with 60% acetone. To each evaporate, 50µl of assay buffer was added followed by 50µl rabbit antibody against oxytocin. After a 60-min preincubation interval, 10µl 125I-labeled tracer (PerkinElmer, Waltham, MA, USA) was added and samples incubated for three days at 4°C. Unbound radioactivity was precipitated by activated charcoal (Sigma-Aldrich, St Louis, MO, USA). Under these conditions, an average of 50% of total counts are bound with <5% non-specific binding. The detection limit of the radioimmunoassay was in the 0.1-0.5 pg/sample range, depending on the age of the tracer, with typical displacements of 20–25% at 2 pg, 60–70% at 8 pg, and 90% at 32 pg of standard neuropeptide. Cross-reactivities with arginine vasopressin, ring moieties and terminal tripeptides of both oxytocin and vasopressin, as well as a wide variety of peptides comprising 3 (alpha-melanocyte-stimulating hormone) up to 41 (corticotropin-releasing factor) amino acids are <0.7% throughout. Intra- and inter-assay coeficients of variation were <10%. Serial dilutions of plasma samples containing high levels of endogenous oxytocin run strictly parallel to the standard curve indicating immuno-identity.

### Questionnaires

State anxiety was measured using the state scale of the German short version of the State-Trait Anxiety Inventory (STAI-s) (Grimm, 2009), which captures acute feelings of tension, apprehension, worry and nervousness. The STAI is a validated and widely used self-rating scale for measuring the severity of anxiety (Spielberger et al., 1983).

Additionally, all participants completed several self-report scales at the end of each visit, three of which were relevant to the present analysis. During the first visit, participants filled out the Beck Depression Inventory (BDI-II) (Beck et al., 1996; Kühner et al., 2006). For the current analysis, we only included items capturing cognitive and affective symptoms, since it has been shown that using self-report measures of depression that include somatic complaints may lead to an overestimation of the prevalence of depression in patients with medical conditions, including substance use disorders (Volk et al., 1993). This procedure is analogous to using the Beck Depression Inventory Fast Screen for medical patients (BDI-FS) (Beck et al., 2000), which has been validated in various clinical populations (Alsaleh et al., 2019; Benedict et al., 2003; Poole et al., 2009).

At the end of the second visit, participants completed the Stress-Reactivity-Scale (SRS) (Schulz et al., 2005), as well as the screening scale of the Trier Inventory for the Assessment of Chronic Stress (TICS) (Schulz & Schlotz, 1999). The SRS assesses a person’s tendency to respond to stressors with immediate, long lasting and intense stress reactions (Schulz et al., 2005). The TICS captures different aspects of subjectively experienced chronic stress, such as work overload or lack of social recognition, over the last three months (Schulz & Schlotz, 1999).

### Statistical analysis

All statistical analyses were performed using R Statistical Software version 4.1.0 (R Core Team, 2021) (for details on used packages, see Supplementary Materials). For each participant and each visit, we calculated the difference in anxiety before and after the task, which was captured in the difference in STAI-s sum scores at T2 vs. T1. Pulse rate recordings were averaged across the resting period as well as the test phase of the stress or control task. Pulse rate change was quantified as the difference between mean pulse rate during the test phase and resting period, for each participant and visit. Cortisol change was calculated using the Area under the curve with respect to increase (AUCi) (Pruessner et al., 2003). Salivary cortisol levels collected at T1 (after the resting period) were chosen as the reference point from which the increase was determined.

Following prior analyses (Martins et al., 2020), we estimated relative and absolute reliability of plasmatic oxytocin levels between the two visits using within-subject coeficients of variation (CV) and intraclass correlations (ICCs). ICCs across the two visits were calculated using a two-way mixed model, single measures, absolute agreement (Koo & Li, 2016). A two sample t-test was performed to compare mean oxytocin levels in participants with AUD and control participants.

The effects of oxytocin on affective and physiological markers of stress were assessed using linear mixed-effect models. Anxiety change, pulse rate change, as well as cortisol change were used as dependent variables. We constructed three models per dependent variable, each containing a random intercept for each participant, as well as task (TSST vs. control task), group (AUD vs. control group) and oxytocin as fixed effects. All models also controlled for gender, age, as well as the order of the stress vs. control task by including them as additional fixed effects. Age and oxytocin were centered on the grand mean. The models differed only in whether and which interaction effects were included: the first model only included main effects, the second model included all main effects and two-way interactions between oxytocin, task and group, and the third model included all main effects, two-way interactions and the three-way interaction of oxytocin × group × task. We used likelihood ratio tests to determine the model that best explained the data for each dependent variable (see Supplementary Table S4 for complete results of the model comparisons). Since the models only differed in their fixed effects structure, they were estimated using maximum likelihood (ML) for the purpose of model comparisons. This is recommended since likelihood values derived from restricted maximum likelihood (REML) depend on the fixed effects in the model (Faraway, 2016). The respective winning model was then re-estimated using REML and significance tests for the model coeficients were performed using Satterthwaite’s approximations for degrees of freedom (Meteyard & Davies, 2020).

## Results

### Good reliability of plasmatic oxytocin measures

Within-subject variation of plasmatic oxytocin measurements is illustrated in **Figure 1B**. The ICC for the total sample was estimated to be 0.79, 95% CI [0.68; 0.87]. For the AUD group, the ICC was slightly lower at 0.71 [0.50; 0.84]. For the control group, it was 0.90 [0.81; 0.95]. The within-subject coeficient of variation (CV) was estimated to be 11.1% for the total sample, and 12.4% and 9.4% for the AUD and control groups, respectively. In general, the findings suggest good reliability of plasma oxytocin levels in our sample.

### No difference in mean oxytocin levels between groups

The distribution of oxytocin levels in the two groups is shown in **Figure 1C**. Since oxytocin levels were relatively stable between the two visits, we used mean values when comparing oxytocin levels in participants with AUD and control participants. Results of a two-sample t-test indicated that there were no significant differences between the groups (*t*(75) = 0.727, *p* = 0.470, *d* = 0.17 [-0.28; 0.61]).

### Oxytocin is associated with affective, but not physiological markers of stress

#### Oxytocin effects on anxiety change

The model containing two-way interactions best explained the change in anxiety (R^2^ marginal = 0.32, R^2^ conditional = 0.42; see also Supplementary Tables S2 and S3). A significant main effect of task suggested that anxiety change was higher in the TSST condition than the control condition (β = 6.96 [5.29, 8.64], *p* < .001). The main effect of oxytocin was not significant (β = 0.08 [-2.90, 3.07]). However, there was a significant oxytocin × group effect (β = 3.61 [0.62, 6.59]), *p* = 0.024), indicating that oxytocin levels were differentially associated with the overall anxiety change in participants with AUD and control participants irrespective of the task. To further explore the oxytocin × group interaction, we conducted post-hoc correlation analyses examining the relationship between mean oxytocin levels and mean change in anxiety separately for each group. The results showed that mean oxytocin levels were positively correlated with mean anxiety change in the AUD group (*r* = 0.274, *p* = 0.087) and negatively correlated in the control group (*r* = −0.274, *p* = 0.101), although both correlations did not reach statistical significance. Further correlation analyses revealed that this effect was mainly driven by differences in the TSST condition (**Figure 2**), where the correlations of oxytocin and anxiety change differed significantly between groups (AUD: *r* = 0.223, controls: *r* = −0.407, *p* = .006, using Fisher’s r-to-Z transformation for the comparison of correlation coeficients). In contrast, the correlations of anxiety change and oxytocin did not differ between the groups under control conditions (AUD: *r* = 0.186, controls: *r* = −0.011, *p* = .403).

**Figure 2:**
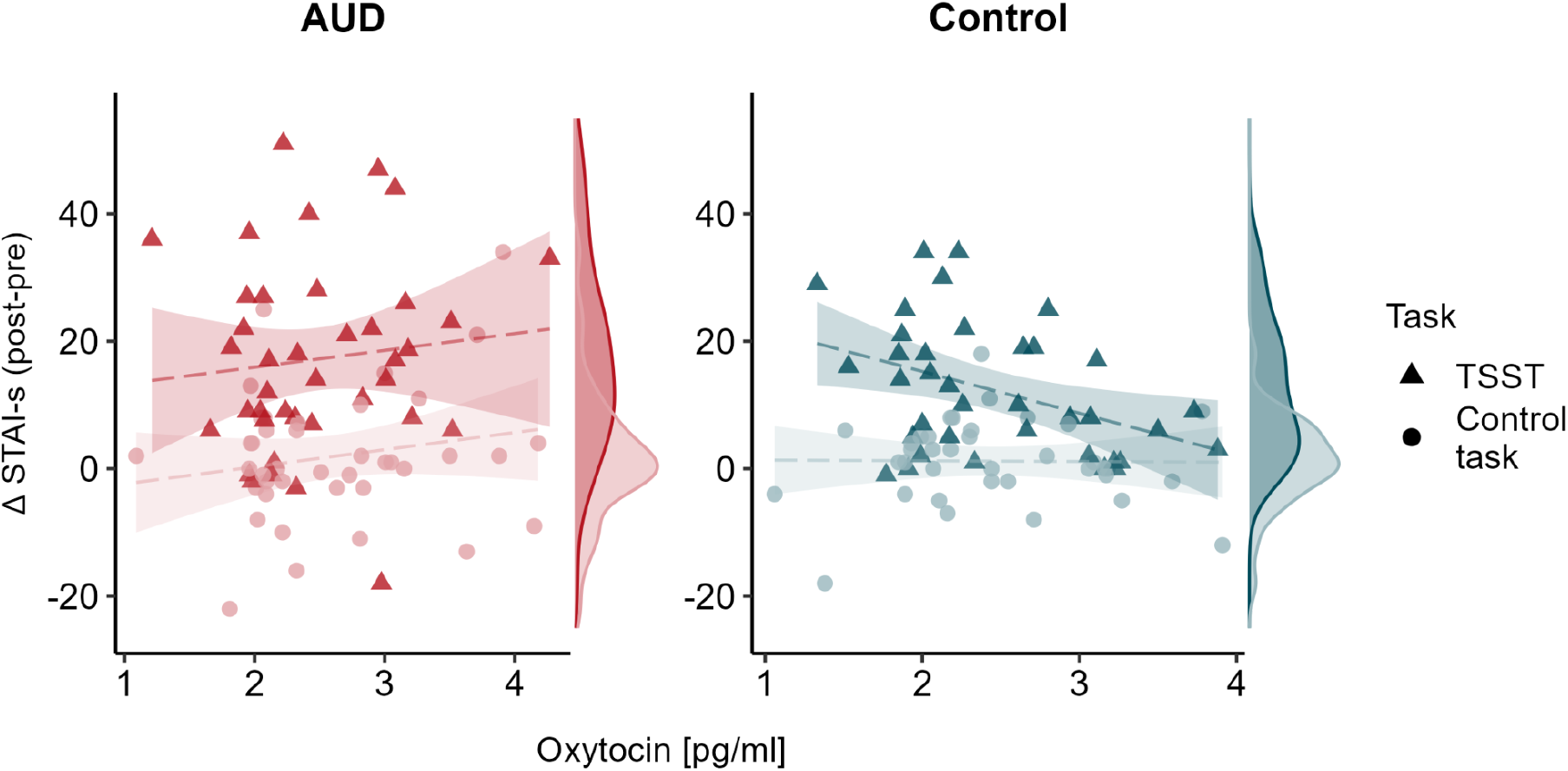
Relationship of oxytocin levels and anxiety change by task and group. Results of a linear mixed model showed a significant group × oxytocin interaction, indicating that mean oxytocin levels were differentially associated with anxiety change in the two groups across the two tasks. This interaction was mainly driven by changes during the TSST condition.

#### No oxytocin effects on pulse rate change

The model including two-way interactions best explained changes in pulse rate (R^2^ marginal = .12, R^2^ conditional = .58; see Supplementary Table S4). A significant main effect of task suggested a greater change in pulse rate in the TSST condition compared to the control condition (β = 2.58 [1.46, 3.73], *p* < .001). Further, there was a significant group × task interaction (β = −1.75 [-2.89, −0.62], *p* = .004). The main effect of oxytocin was not significant (β = −0.20 [-3.09, 2.68]) and there were no significant interactions of oxytocin with task (β = −1.01 [-2.79, 0.76]) or group (β = −1.16 [-4.08, 1.79]), indicating that oxytocin levels were not associated with the change of pulse rate over the course of the tasks.

#### No oxytocin effects on cortisol release

The model including two-way interactions best explained the increase in cortisol levels (R^2^ marginal = 0.21, R^2^ conditional = 0.43; see Supplementary Table S5). Again, a significant main effect of task suggested that the increase in cortisol levels was higher in the TSST condition than the control condition (β = 127.5 [86.0, 169.0], *p* < .001). The analysis also yielded a significant group × task interaction (β = -57.8 [-99.5, −16.2], *p* = .009). The main effect of oxytocin was not significant (β = -32.4 [-115.7, 50.8]) and there were no significant interactions of oxytocin with task (β = −1.3 [-65.8, 63.7]) or group (β = 67.1 [-16.3, 150.6]). The results suggest that there was no relationship between plasmatic oxytocin levels and cortisol release over the course of the visits.

#### Exploratory analyses – altered association of oxytocin and depressive symptoms in AUD

Oxytocin levels seem to be associated with affective, but not physiological stress responses in our sample. To explore whether oxytocin levels were differentially associated with other measures of affective experiences, we performed three additional linear regressions examining the relationship between oxytocin and depressive symptoms, trait stress reactivity and perceived chronic stress. All models included mean oxytocin levels, group, as well as their interaction term as predictors. BDI-FS, SRS, as well as TICS screening scores were used as outcome variables, respectively. There was a significant main effect of oxytocin on BDI-FS scores (β = 1.37 [0.22, 2.52], *p* = 0.020), indicating an overall increase in affective and cognitive depressive symptoms with increasing plasmatic oxytocin. However, the group × oxytocin interaction was also significant (β= −2.15 [-3.81, −0.49], *p* = 0.012), suggesting that oxytocin levels were differentially associated with depressive symptoms in the two groups. Post-hoc correlation analyses showed that oxytocin levels were positively associated with depressive symptoms in the AUD group (Spearman ρ = 0.29, *p* = 0.073), and negatively associated with depressive symptoms in the control group (ρ = −0.40, *p* = 0.014; **Figure 3**). Group, oxytocin or the interaction term did not significantly predict stress reactivity or perceived chronic stress (Supplementary Table S6).

**Figure 3:**
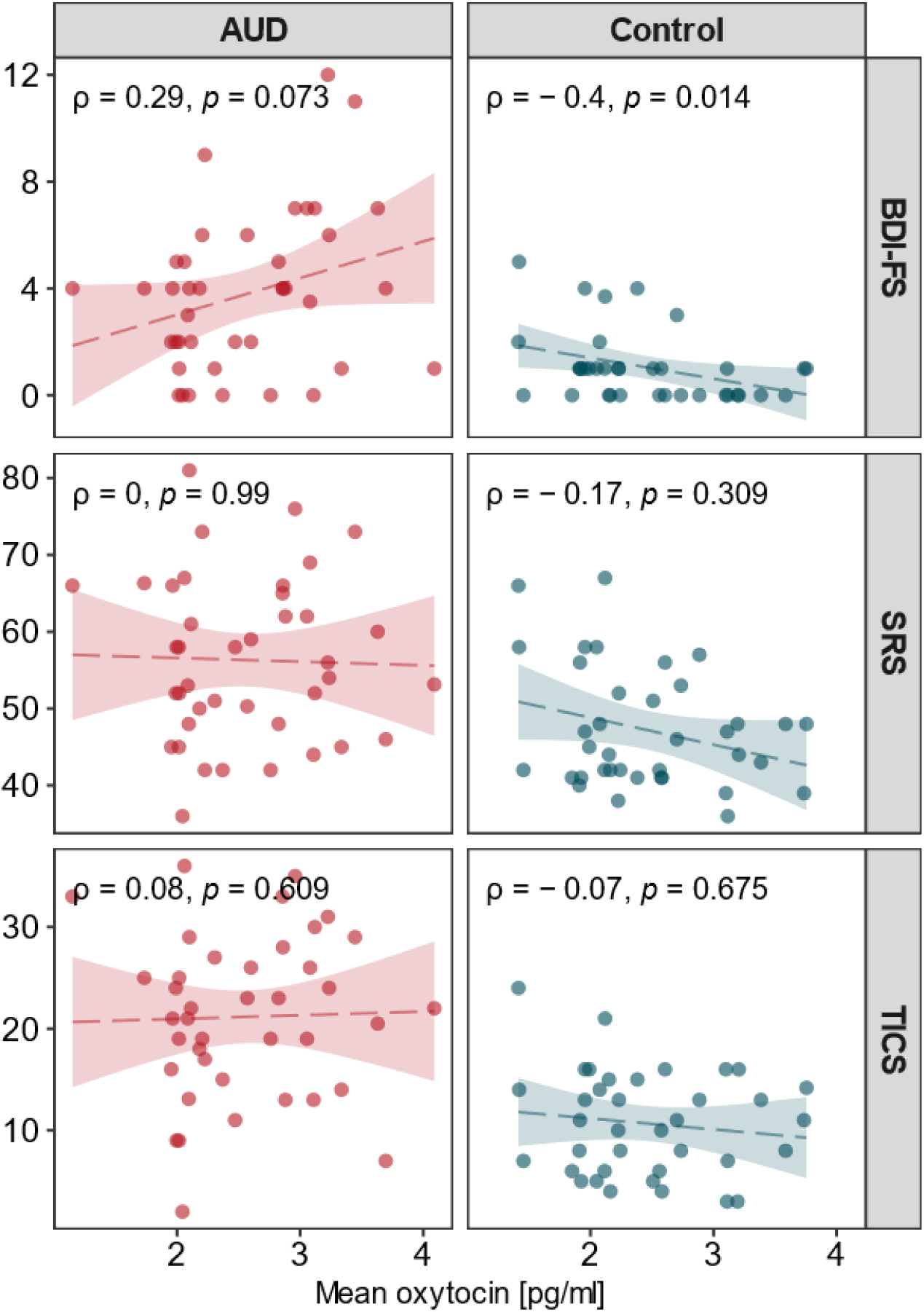
Correlations of mean oxytocin values with non-somatic symptoms of depression (BDI-FS), trait stress reactivity (SRS) and perceived chronic stress (TICS). The association between oxytocin levels and depressive symptoms was significantly different between the AUD and control groups, as indicated by a significant group × oxytocin interaction in a linear regression analysis.

## Discussion

Although it has been proposed that oxytocin plays a role in the development and maintenance of addiction through interactions with stress pathways, direct evidence of a relationship between endogenous oxytocin levels and stress reactivity in AUD is sparse. In this study we investigated the relationship between baseline plasma oxytocin levels and responses to stress in individuals with AUD. Our main findings suggest an altered association of oxytocin and affective, but not physiological, stress responses in individuals with AUD. While participants with AUD showed a positive relationship between plasmatic oxytocin levels and stress-induced anxiety change, the opposite relationship was found in control participants. A similar pattern of results was found for affective and cognitive symptoms of depression in exploratory analyses, indicating that higher oxytocin levels were associated with higher depression levels in individuals with AUD, but lower depression levels in control participants.

In healthy individuals, previous studies have shown that higher endogenous oxytocin levels correlate with reduced anxiety responses (Carson et al., 2015; Heinrichs et al., 2003; Weisman et al., 2013), a pattern that we also replicate in our control sample. However, in subjects with AUD, oxytocin appears not to have the same anxiolytic properties under stressful conditions. This finding might explain results from clinical intranasal oxytocin trials that were conducted expecting the same anxiolytic effects as in healthy participants, but showed diverging effects in psychiatric patients. For example, in a study examining oxytocin effects in individuals with depression, oxytocin led to increased anxiety levels over the course of a psychotherapy session, contrary to the authors’ hypothesis (MacDonald et al., 2013). Assuming that affective responses to acute stress mediate addiction-related behaviors, this positive correlation of oxytocin levels and increased anxiety suggests that individuals with AUD might not benefit from oxytocin administrations to upregulate the oxytocin system. Indeed, recent findings from randomized controlled trials showed that intranasal oxytocin did not reduce alcohol craving (Melby et al., 2021; Stauffer et al., 2019) or prevent relapse in patients with AUD after detoxification (Melby et al., 2021). It is worth noting, however, that findings from clinical oxytocin trials have been heterogenous, and studies on oxytocin effects in humans with AUD are still scarce (Mellentin et al., 2023).

Nevertheless, our results are in line with the notion that oxytocin effects are modulated by contextual and individual differences (Bartz et al., 2011; Olff et al., 2013) and highlight the importance of considering these differences when making predictions about the efficacy of oxytocin interventions.

While oxytocin levels were differentially correlated with changes in anxiety after stress in our sample, we found no significant relationship between oxytocin and pulse rate change or salivary cortisol increase in either group. At first glance, this finding seems to contradict evidence from animal and human studies showing that oxytocin administration has stress-buffering effects on the HPA-axis (Cardoso et al., 2013; Heinrichs et al., 2003; Linnen et al., 2012; Neumann et al., 2000) as well as the autonomic nervous system (Wsol et al., 2009). However, only few studies have examined associations of *endogenous* oxytocin and physiological stress responses, and the evidence from these studies has been heterogeneous. For example, higher overall plasmatic oxytocin levels were linked to a stronger decline in heart rate during a cold pressor task, but not during a public speaking task in postpartum mothers (Grewen & Light, 2011). Another study showed that there was no correlation of oxytocin and cortisol increase in the TSST in older women (Taylor et al., 2006). Thus, our results align with previous findings on the relationship between plasma oxytocin levels and physiological changes triggered by psychosocial stress, which appear to differ from studies involving oxytocin administration.

Contrary to previous studies (Lenz et al., 2021; Marchesi et al., 1997), we did not find evidence for elevated baseline oxytocin levels in patients with AUD compared to healthy control participants. One possible explanation for this might lie in the time interval between withdrawal and study participation. In individuals with AUD, it has been shown that oxytocin blood concentrations continuously decrease during the first days of abstinence, with one study reporting no significant differences compared to a healthy control group after around five days (Lenz et al., 2021). Another study similarly reported a significant decrease of blood oxytocin levels after day 4, although oxytocin levels were still elevated compared to controls after one month of abstinence (Marchesi et al., 1997). However, since the sample size of the latter study was considerably smaller (Marchesi et al., 1997: *N* = 22) than of the former (Lenz et al., 2021: *N* = 440), it is reasonable to expect that oxytocin levels in participants with and without AUD are already comparable within the first few weeks of abstinence. Since we included patients who had been abstinent for nine to 39 days at the time of study participation, it is thus not surprising that we found similar oxytocin levels in the two groups. Together with previous evidence, this suggests that elevated blood oxytocin levels in AUD are a reversible consequence of excessive alcohol consumption, rather than a risk factor for addiction.

Our reliability analysis revealed that baseline plasmatic oxytocin measures were stable within individuals between the two visits. On a descriptive level, reliability measures tended to be lower within the AUD group, which might be explained by the aforementioned decline of oxytocin levels in the first weeks of abstinence (Lenz et al., 2021; Marchesi et al., 1997).

However, even within the AUD group, reliability measures were good and did not substantially differ from the control group. This finding stands in contrast to the only previous study that evaluated the reliability of peripheral oxytocin concentrations and observed that salivary and plasmatic oxytocin were not stable over time, concluding that single measurements are not valid trait markers of the oxytocin system in humans (Martins et al., 2020). It is important to note that our methods to quantify oxytocin concentrations and analyze their reliability were similar or even identical to those reported in Martins et al. (2020): (1) we collected blood samples at roughly the same time in the morning to avoid confounding effects of daytime fluctuations of oxytocin, (2) samples were processed immediately after their collection, (3) plasma samples were extracted prior to quantification, (4) quantification was carried out using radioimmunoassay in the same laboratory, thus following the same standard procedures, and (5) between-visits reliability was estimated using ICC and CV to ensure comparability of the results. It is therefore unclear why we found different results. In any case, our results do not support the conclusion that baseline plasma oxytocin levels are generally unstable under typical laboratory conditions.

A strength of our study is that we were the first to empirically investigate associations of endogenous oxytocin system functioning and stress reactivity in AUD, which is an important step towards the understanding of addiction and the development of targeted pharmacological interventions. However, when interpreting the present results, one may consider the following aspects. First, plasma oxytocin, that is, peripheral oxytocin concentrations, were analyzed, although oxytocin is assumed to exert effects on stress responses mainly via central release (Takayanagi & Onaka, 2022). While plasmatic oxytocin is regularly used in human studies since collecting blood samples is less invasive than collecting centrally circulating fluids, there have been doubts on whether peripheral oxytocin concentrations are a good approximation for central oxytocin concentrations, especially under baseline conditions (Valstad et al., 2017). Second, oxytocin effects are often found to be moderated by interindividual differences, for example in early chronic stress experiences, bonding behavior, social support and variation in the oxytocin or oxytocin receptor gene (Olff et al., 2013), which we did not investigate here. Further, while we did include women in our study, our sample was predominantly male. Since there have been various reports of sex-dependent oxytocin effects on stress responses (Kubzansky et al., 2012; Love, 2018), our conclusions may be limited to men only. Third, it is important to note that this was a correlational study, which means that our findings cannot be used to make claims about a potential causal role of oxytocin-system functioning in AUD. Whether altered associations of oxytocin and affective stress responses represent a risk factor for AUD or a consequence of prolonged alcohol consumption thus remains an open question.

To conclude, our findings suggest that endogenous oxytocin levels may not be altered at baseline in individuals with AUD during abstinence, but rather are differentially related to affective responses to psychosocial stress. Our findings challenge the notion that enhancing the endogenous oxytocin system is universally beneficial in treating addiction. Instead, if affective responses to stress drive addiction-related behaviors like craving and relapse, administering exogenous oxytocin might not help individuals with AUD, at least during the early stages of abstinence. While it is likely that elevated baseline oxytocin levels are a reversible consequence of prolonged alcohol consumption, future studies should address whether altered associations of oxytocin and stress-related anxiety persist even in long-term abstinence.

## Data Availability

Data and code of this study are openly available at

https://osf.io/gjs5h/

## Acknowledgements

This work was supported by the Else Kröner-Fresenius-Stiftung (to LR, Grant No. 2018_A26). AVM received funding by the Department of Medicine at the University of Lübeck, CS08-2023. YS was funded by the Studienstiftung des Deutschen Volkes.

We thank Julie Forster, Jovana Lehmann-Grube and Emil Zillikens for their help in data collection.

## Disclosures

The authors report no biomedical financial interests or potential conflicts of interest. Funding was acquired by LR. LR, JV, SK, and FMP designed the study. YS, JS, JV, AS, OV, JG, and KJ recruited the participants and acquired the data. AVM, LR, SK, and FMP analyzed data. AVM drafted the manuscript. All authors critically reviewed the manuscript and approved the final version.

Data and code of this study are openly available at: https://osf.io/gjs5h/

