## Supplementary Materials for "Altered association of plasmatic oxytocin with affective stress response in alcohol use disorder"

### Supplementary Materials for: Altered association of plasmatic oxytocin with affective stress responses in alcohol use disorder

Mayer, A.V., Schwarze, Y., Stierand, J., Voges, J., Schröder, A., von der Gablentz, J., Junghanns, K., Voß, O., Krach, S., Paulus, F.M., & Rademacher, L.

#### Inclusion and exclusion criteria

General inclusion criteria were an age between 18 and 60 years and sufficient German language skills. Participants were excluded if they (1) suffered from any neurological disease, traumatic brain injuries, acute inflammatory disease or chronic inflammatory bowel disease, (2) were taking medication that exerted a direct influence on the HPA axis, in particular beta-blockers or medication containing hydrocortisone. Since parts of this study were carried out using magnetic resonance imaging (MRI), subjects were also required to be free of any metallic implants such as pacemakers, defibrillators or copper IUDs, as well as metallic splinters or shrapnel. Other MRI contraindications included large tattoos or permanent make-up, claustrophobia, and pregnancy.

Participants with alcohol use disorder (AUD) were additionally required to meet the criteria for moderate to severe AUD according to the Diagnostic and Statistical Manual of Mental Disorders fifth edition (DSM-5) and to be abstinent for at least 9 days at the time of study participation. Participants with AUD were excluded if they (1) had a history of substance abuse or dependency other than alcohol or nicotine within the last 12 months, (2) were diagnosed with major depression, social anxiety disorder, antisocial or borderline personality disorder, (3) were in an acute state of psychosis or suicidality. Control participants were excluded if they (1) suffered from any current or past psychiatric disorders, (2) scored ≥8 on the Alcohol Use Disorder Identification Test (AUDIT) (Bohn et al., 1995), (3) had any first-degree relatives with alcohol use disorder, (4) had consumed drugs (except for alcohol, nicotine, and irregular use of cannabis) in the past year. All inclusion and exclusion criteria were checked by telephone or in a personal interview before participants were included in the study.

#### Changes in study protocol due to SARS-CoV-2 pandemic

Due to the SARS-CoV-2 pandemic, data collection was temporarily halted between March 2020 and October 2021. The study was continued as soon as the risk for infection was sufficiently reduced by available vaccinations and testing. Forty-nine participants were enrolled before the pandemic, and thirty-one participants were enrolled during the pandemic. Following the pause, several procedural adjustments were made. For instance, participants underwent a rapid COVID-19 test upon arrival, and face masks were worn throughout the visits. Additionally, before the pandemic, blood samples were collected at four time points using indwelling venous catheters: ten minutes after arrival (T0), immediately after the stress or control task (T2), after MRI measurements (T4) and at the end of each visit (T5). During the pandemic, we decided to reduce the number of blood draws. Blood was now only collected at the beginning (T0) and end (T5) of each study visit using venipuncture.

##### MRI experiments

The present analysis is part of a larger fMRI study investigating the acute behavioral, physiological, and neural responses to social stress in AUD, involving two MRI experiments for participants. Both experiments examined brain function associated with social vs. non-social stimuli, which might be altered after stress in individuals with AUD. The first experiment was a modified version of the monetary and social incentive delay (MID/SID) paradigm (Knutson et al., 2001; Rademacher et al., 2010; Spreckelmeyer et al., 2009), which is a well-established task to study participants' neural responses to anticipated rewards. During the task, participants respond as quickly as possible to a target stimulus after visual cues indicate a potential reward, with fast responses earning either social rewards (smiling faces) or monetary rewards (small amounts of money). This task is known to activate the dopaminergic reward system, which has been associated with addiction and stress. The second experiment involved a novel paradigm aimed at exploring neural and behavioral responses to images of social situations. Participants viewed images of both social and non-social scenes with differing numbers of people and were then asked to rate how much they would like to be in the depicted situation. Results of the fMRI analyses will be reported elsewhere.

##### Analysis software

We used R version 4.1.0 (R Core Team, 2021) and the following R packages: afex v. 1.3.0 (Singmann et al., 2023), arsenal v. 3.6.3 (Heinzen et al., 2021), cocor v. 1.1.3 (Diedenhofen & Musch, 2015), DescTools v. 0.99.48 (Signorell, 2023), effectsize v. 0.8.5 (Ben-Shachar et al., 2020), ggExtra v. 0.10.1 (Attali & Baker, 2023), ggpubr v. 0.6.0 (Kassambara, 2023), ggrain v. 0.0.3 (Allen et al., 2021), irr v. 0.84.1 (Gamer et al., 2019), lme4 v. 1.1.28 (Bates et al., 2015), mlmtools v. 1.0.2 (Jamison et al., 2022), and sjPlot v. 2.8.16 (Lüdecke, 2024).

#### Supplementary Tables

**Table S1. Summary of affective and physiological stress measures per task and group.**

| | AUD group (N=40) | | Control group (N=37) | |
|---|---|---|---|---|
| | **Control task** | **TSST** | **Control task** | **TSST** |
| **Anxiety pre task [STAI-S]** | | | | |
| Mean (SD) | 24.34 (10.12) | 23.15 (9.96) | 23.57 (8.27) | 22.49 (8.40) |
| Range | 12 - 57 | 10 - 52 | 10 - 41 | 10 - 46 |
| **Anxiety post task [STAI-S]** | | | | |
| Mean (SD) | 26.28 (10.39) | 41.48 (13.89) | 24.76 (7.69) | 34.70 (12.28) |
| Range | 12 - 50 | 12 - 73 | 10 - 39 | 10 - 59 |
| **Cortisol T0 [nmol/l]** | | | | |
| Mean (SD) | 9.22 (5.79) | 8.98 (5.43) | 8.22 (4.89) | 8.68 (5.49) |
| Range | 2.38 - 23.39 | 2.21 - 23.17 | 1.74 - 27.39 | 0.99 - 24.28 |
| **Cortisol T1 [nmol/l]** | | | | |
| Mean (SD) | 6.76 (5.97) | 5.95 (4.85) | 8.35 (7.20) | 8.64 (8.72) |
| Range | 0.95 - 38.50 | 1.35 - 29.75 | 1.54 - 34.14 | 1.28 - 37.83 |
| **Cortisol T2 [nmol/l]** | | | | |
| Mean (SD) | 5.01 (3.74) | 5.36 (3.34) | 6.49 (5.84) | 10.20 (7.52) |
| Range | 1.02 - 23.53 | 1.30 - 18.80 | 1.23 - 23.68 | 1.82 - 27.80 |
| **Cortisol T3 [nmol/l]** | | | | |
| N-Miss | 1 | 0 | 0 | 0 |
| Mean (SD) | 4.05 (2.63) | 6.19 (3.29) | 5.24 (3.57) | 13.28 (8.40) |
| Range | 1.13 - 13.94 | 1.47 - 14.81 | 1.12 - 13.70 | 1.63 - 32.54 |
| **Cortisol T4 [nmol/l]** | | | | |
| Mean (SD) | 4.19 (4.20) | 4.26 (2.74) | 5.00 (5.20) | 6.52 (3.95) |
| Range | 1.35 - 28.05 | 0.83 - 17.20 | 0.95 - 32.85 | 1.07 - 19.59 |
| **Cortisol increase from T1 [AUCi]** | | | | |
| N-Miss | 1 | 0 | 0 | 0 |
| Mean (SD) | 165 (219) | 300 (216) | 155 (220) | 524 (494) |
| Range | -417 - 956 | -92 - 1001 | -767- 636 | -366 - 2027 |
| **Pulse rate pre task [bpm]** | | | | |
| N-Miss | 0 | 0 | 1 | 0 |
| Mean (SD) | 81.29 (12.13) | 79.82 (13.18) | 71.16 (11.70) | 71.91 (12.52) |
| Range | 56.12 - 111.03 | 51.19 - 104.43 | 48.76 - 100.13 | 48.43 - 105.80 |
| **Pulse rate during task [bpm]** | | | | |
| N-Miss | 0 | 0 | 1 | 1 |
| Mean (SD) | 92.34 (13.44) | 92.59 (19.07) | 79.64 (12.20) | 89.98 (17.75) |
| Range | 69.94 - 124.79 | 54.99 - 130.30 | 55.42 - 103.92 | 53.22 - 135.15 |

*Note.* STAI-S: State-Trait Anxiety Inventory - State measure (Laux et al., 1981). Higher STAI-S values indicate higher levels of subjectively perceived nervousness, tension, and worry. AUCi: area under the curve with respect to increase (Pruessner et al., 2003), calculated between T1 and T4. Higher AUCi indicates a stronger increase in cortisol levels between T1 and T4. Negative values indicate a decrease of cortisol levels between T1 and T4.

**Table S2: Model comparisons of linear mixed-effect models.**

| DV | Model name | Model fit | | | LRT against nested | | |
|---|---|---|---|---|---|---|---|
| | | **AIC** | **BIC** | **LL** | **df** | **$\chi^2$** | ***p*** |
| Anxiety change | main effects model | 1202.9 | 1230.2 | -592.4 | | | |
| | **two-way interactions model** | 1200.4 | 1236.8 | -588.2 | 3 | 8.51 | 0.037 |
| | three-way interaction model | 1199.6 | 1239.1 | -586.8 | 1 | 2.81 | 0.094 |
| Pulse rate change | main effects model | 1135.5 | 1162.7 | -558.8 | | | |
| | **two-way interactions model** | 1130.7 | 1167.0 | -553.4 | 3 | 10.79 | 0.013 |
| | three-way interaction model | 1132.7 | 1172.0 | -553.4 | 1 | 0.00 | 0.994 |
| Cortisol change | main effects model | 2200.1 | 2227.4 | -1091.1 | | | |
| | **two-way interactions model** | 2196.1 | 2232.4 | -1086.0 | 3 | 10.09 | 0.018 |
| | three-way interaction model | 2197.9 | 2237.3 | -1085.9 | 1 | 0.19 | 0.664 |

*Note*. We constructed three models per dependent variable, each containing a random intercept for each participant, as well as task (TSST vs. control task), group (AUD vs. control group) and oxytocin as fixed effects. The models differed only in whether and which interaction effects were included: the first model only included main effects, the second model included all main effects and two-way interactions between oxytocin, task and group, and the third model included all main effects, two-way interactions and the three-way interaction of oxytocin × group × task. All models were estimated using maximum likelihood (ML) for the purpose of model comparisons. The respective winning models are highlighted in bold. DV: dependent variable, LRT = likelihood ratio test, AIC = Akaike information criterion, BIC = Bayesian information criterion, LL = log likelihood.

**Table S3: Oxytocin and anxiety change: results of linear mixed-effects model.**

| **Fixed effects** | | | | | | |
|---|---|---|---|---|---|---|
| | Beta | SE | 95% CI | | *t* | *p* |
| | | | lower | upper | | |
| Intercept | 5.77 | 2.24 | 1.52 | 10.01 | 2.57 | 0.012 |
| Group | 1.71 | 0.99 | -0.18 | 3.59 | 1.72 | 0.090 |
| Age | 0.07 | 0.12 | -0.15 | 0.29 | 0.57 | 0.571 |
| Gender | -2.75 | 2.24 | -6.99 | 1.49 | -1.23 | 0.223 |
| Task | 6.96 | 0.86 | 5.29 | 8.64 | 8.10 | **< 0.001** |
| Order | 1.23 | 1.02 | -0.71 | 3.16 | 1.20 | 0.233 |
| Oxytocin | 0.08 | 1.57 | -2.90 | 3.07 | 0.05 | 0.959 |
| Group × Oxytocin | 3.61 | 1.57 | 0.62 | 6.59 | 2.29 | **0.024** |
| Group × Task | 1.46 | 0.86 | -0.22 | 3.13 | 1.69 | 0.095 |
| Task × Oxytocin | -0.71 | 1.34 | -3.25 | 2.01 | -0.53 | 0.595 |

| **Random effects** | | |
|---|---|---|
| | Variance | SD |
| Participant (Intercept) | 18.2 | 4.3 |

| **Model fit** | | |
|---|---|---|
| $R^2$ | Marginal | Conditional |
| | 0.32 | 0.42 |

*Note.* P-values for fixed effects have been calculated using Satterthwaite's approximations.
Model equation: anxiety difference ~ (1|id)+ group + age + gender + task + order + oxytocin + oxytocin*group + group*task + oxytocin*task

**Table S4: Oxytocin and pulse rate change: results of linear mixed-effects model.**

| **Fixed effects** | | | | | | |
|---|---|---|---|---|---|---|
| | Beta | SE | 95% CI lower | 95% CI upper | *t* | *p* |
| Intercept | 12.97 | 2.41 | 8.39 | 17.55 | 5.38 | <0.001 |
| Group | -0.81 | 1.04 | -2.79 | 1.16 | -0.78 | 0.438 |
| Age | -0.11 | 0.12 | -0.34 | 0.11 | -0.95 | 0.347 |
| Gender | 0.33 | 2.41 | -4.25 | 4.91 | 0.14 | 0.891 |
| Task | 2.58 | 0.58 | 1.46 | 3.73 | 4.44 | **< 0.001** |
| Order | -1.49 | 1.06 | -3.51 | 0.52 | -1.41 | 0.164 |
| Oxytocin | -0.20 | 1.52 | -3.09 | 2.68 | -0.13 | 0.895 |
| Group × Oxytocin | -1.16 | 1.52 | -4.08 | 1.79 | -0.76 | 0.447 |
| Group × Task | -1.75 | 0.58 | -2.89 | -0.62 | -2.99 | **0.004** |
| Task × Oxytocin | -1.01 | 0.92 | -2.79 | 0.76 | -1.10 | 0.274 |

| **Random effects** | | |
|---|---|---|
| | Variance | SD |
| Participant (Intercept) | 56.39 | 7.51 |

| **Model fit** | | |
|---|---|---|
| $R^2$ | Marginal | Conditional |
| | 0.12 | 0.58 |

Note. *P*-values for fixed effects have been calculated using Satterthwaite's approximations.
Model equation: pulse rate difference ~ (1|id)+ group + age + gender + task + order + oxytocin + oxytocin*group + group*task + oxytocin*task

**Table S5: Oxytocin and cortisol increase: results of linear mixed-effects model.**

| **Fixed effects** | | | | | | |
|---|---|---|---|---|---|---|
| | Beta | SE | 95% CI lower | 95% CI upper | *t* | *p* |
| Intercept | 208.2 | 63.9 | 87.1 | 329.3 | 3.26 | 0.002 |
| Group | -52.5 | 28.4 | -106.4 | 1.3 | -1.85 | 0.069 |
| Age | -0.8 | 3.3 | -7.0 | 5.5 | -0.23 | 0.818 |
| Gender | -82.7 | 63.8 | -203.6 | 38.2 | -1.30 | 0.199 |
| Task | 127.5 | 21.4 | 86.0 | 169.0 | 5.97 | **< 0.001** |
| Order | 8.0 | 29.1 | -47.1 | 63.1 | 0.28 | 0.784 |
| Oxytocin | -32.4 | 43.9 | -115.7 | 50.8 | -0.74 | 0.461 |
| Group × Oxytocin | 67.1 | 44.0 | -16.3 | 150.6 | 1.53 | 0.130 |
| Group × Task | -57.8 | 21.4 | -99.5 | -16.2 | -2.70 | **0.009** |
| Task × Oxytocin | -1.3 | 33.5 | -65.8 | 63.7 | -0.04 | 0.968 |

| **Random effects** | | |
|---|---|---|
| | Variance | SD |
| Participant (Intercept) | 26017 | 161 |

| **Model fit** | | |
|---|---|---|
| $R^2$ | Marginal | Conditional |
| | 0.21 | 0.43 |

Note. *P*-values for fixed effects have been calculated using Satterthwaite's approximations.
Model equation: AUCi ~ (1|id)+ group + age + gender + task + order + oxytocin + oxytocin*group + group*task + oxytocin*task

**Table S6: Oxytocin, stress and depression: results of linear regression analyses.**

| | Depression [BDI-FS] | | | Stress reactivity [SRS] | | | Chronic stress [TICS] | | |
|---|---|---|---|---|---|---|---|---|---|
| Predictors | β | *CI* | *p* | β | *CI* | *p* | β | *CI* | *p* |
| Intercept | 0.28 | -2.74 – 3.30 | 0.854 | 57.55 | 45.07 – 70.04 | **<0.001** | 20.24 | 11.11 – 29.37 | **<0.001** |
| Mean oxytocin | 1.37 | 0.22 – 2.52 | **0.020** | -0.48 | -5.22 – 4.26 | 0.840 | 0.36 | -3.10 – 3.83 | 0.836 |
| Group | 2.68 | -1.59 – 6.96 | 0.215 | -1.68 | -19.37 – 16.01 | 0.850 | -6.92 | -19.86 – 6.02 | 0.290 |
| Mean oxytocin × Group | -2.15 | -3.81 – -0.49 | **0.012** | -3.04 | -9.89 – 3.81 | 0.379 | -1.44 | -6.45 – 3.58 | 0.570 |
| Observations | 76 | | | 77 | | | 77 | | |
| $R^2$ / $R^2$ adjusted | 0.328 / 0.300 | | | 0.217 / 0.185 | | | 0.384 / 0.358 | | |

*Note.* BDI-FS: Beck Depression Inventory Fast Screen (Beck et al., 2000), SRS: Stress-Reactivity-Scale (Schulz et al., 2005), TICS: screening scale of the Trier Inventory for the Assessment of Chronic Stress (Schulz & Schlotz, 1999). Significant effects are highlighted in bold.
